# Cost Comparison of Fixed Dose Combination vs. Monotherapy Antihypertensive Therapies: A Medicare Part D Analysis (2013 to 2022)

**DOI:** 10.64898/2025.12.01.25341382

**Authors:** Nabeel Sami, Dhara Rana, Ariel Milshteyn, Niloufar Novin, Aparna Dintakurti

## Abstract

Fixed-dose combination (FDC) antihypertensive therapies are widely used to improve adherence and simplify hypertension management, but their long-term cost patterns compared with prescribing individual component drugs remain unclear. We analyzed national Medicare Part D Prescription Drug Event data from 2013 to 2022 to compare the average cost per claim and per beneficiary for six two-drug FDC antihypertensive therapies and the summed costs of their individual components. Analyses were limited to years with complete data for both the combination and its components, and ten-year averages were calculated. Three of the six FDC therapies were less expensive than prescribing their components separately. The greatest savings were seen with valsartan/hydrochlorothiazide ($1 per claim, $174 per beneficiary), lisinopril/hydrochlorothiazide ($4 per claim, $20 per beneficiary), and atenolol/chlorthalidone ($12 per claim, $44 per beneficiary). Amlodipine/valsartan ($63 per claim, $115 per beneficiary), telmisartan/amlodipine ($105 per claim, $386 per beneficiary), and irbesartan/hydrochlorothiazide ($3 per claim, $12 per beneficiary) were more expensive. This national Medicare analysis found that several fixed-dose combination antihypertensive therapies cost less than prescribing their individual components, while others remain more expensive. These results highlight clear differences in drug pricing that may help guide value-based prescribing and formulary decisions in hypertension management.

## Introduction

Fixed-dose combination (FDC) antihypertensive therapies are widely used to improve adherence, reduce pill burden, and simplify blood pressure management [1–3]. Current U.S. guidelines, including the 2017 ACC/AHA Hypertension Guideline, recommend initiating combination therapy in patients with stage 2 hypertension or those not achieving target blood pressure on monotherapy [3]. Randomized trials such as UMPIRE and TRIUMPH have shown that FDCs improve adherence and blood pressure control compared with separate agents [1,2].

Although these regimens offer practical and clinical benefits, their cost relative to prescribing individual component drugs has not been well characterized in real-world practice. Previous modeling and trial-based analyses have suggested that low-dose combinations may be cost-effective [4], but such projections may not reflect contemporary market pricing or formulary dynamics. This question is especially relevant for Medicare Part D, which covers outpatient prescriptions for more than 40 million older adults in the United States and provides insight into national prescribing and cost trends.

The purpose of this study was to describe ten-year national cost trends for six commonly prescribed two-drug fixed-dose combination antihypertensive therapies compared with the summed costs of their component monotherapies using Medicare Part D data from 2013 through 2022.

## Materials and Methods

We used publicly available Medicare Part D Prescription Drug Event data from 2013 through 2022. Medicare Part D covers more than 40 million beneficiaries annually and provides a nationally representative sample of the U.S. older adult population. The dataset includes national-level information on outpatient prescription drug claims for Medicare beneficiaries, including total annual spending, number of claims, and number of unique beneficiaries for each drug. This broad coverage allows for evaluation of real-world prescribing and cost patterns in hypertension management.

Six fixed-dose combination (FDC) antihypertensive therapies were included if they: (1) were consistently reported in the dataset across all ten years, (2) contained exactly two active pharmacologic ingredients, and (3) had publicly available data for both monotherapy components. The selected combinations were clinically relevant, guideline-endorsed, and widely prescribed, representing multiple drug classes. The therapies analyzed were valsartan/hydrochlorothiazide, telmisartan/amlodipine, lisinopril/hydrochlorothiazide, irbesartan/hydrochlorothiazide, amlodipine/valsartan, and atenolol/chlorthalidone. Other fixed-dose combinations were excluded because complete annual data were not available for both the combination and its component drugs across the entire study period.

The primary measures were cost per claim and cost per beneficiary for each FDC and its corresponding component drugs. Cost per claim was calculated by dividing total annual spending by the number of claims, while cost per beneficiary was calculated by dividing total spending by the number of unique beneficiaries. For each year of the study period, the FDC’s cost metrics were compared with the summed costs of its individual components. Analyses were descriptive and limited to years in which complete data were available for both the combination and its monotherapy components. For each drug pair, ten-year average values were calculated, and the difference between averages represented the mean cost difference, reflecting whether the FDC was more or less expensive than its separate agents.

## Results

For the study period from 2013 through 2022, three of the six fixed-dose combination (FDC) antihypertensive therapies were more expensive than their individual component drugs. The combinations with higher costs were amlodipine/valsartan (average increase: $63 per claim and $115 per beneficiary), telmisartan/amlodipine (average increase: $105 per claim and $386 per beneficiary), and irbesartan/hydrochlorothiazide (average increase: $3 per claim and $12 per beneficiary).

In contrast, three FDC therapies showed lower ten-year average costs per claim and per beneficiary compared with their component monotherapies. Cost reductions were observed for valsartan/hydrochlorothiazide (average decrease: $1 per claim and $174 per beneficiary), lisinopril/hydrochlorothiazide (average decrease: $4 per claim and $20 per beneficiary), and atenolol/chlorthalidone (average decrease: $12 per claim and $44 per beneficiary).

Lisinopril/hydrochlorothiazide was by far the most frequently prescribed FDC, with 137.6 million claims and 32.9 million beneficiaries during the study period. The average cost per claim and per beneficiary for lisinopril/hydrochlorothiazide ($9 per claim, $37 per beneficiary) was comparable to lisinopril monotherapy alone ($7 per claim, $34 per beneficiary). Among the remaining combinations, valsartan/hydrochlorothiazide had 50.7 million claims and atenolol/chlorthalidone had 11.8 million claims, representing the next most frequently utilized therapies.

A detailed comparison of ten-year average costs per claim and per beneficiary for each fixed-dose combination and its monotherapy components is summarized in **Table□1**.

**Table 1.**
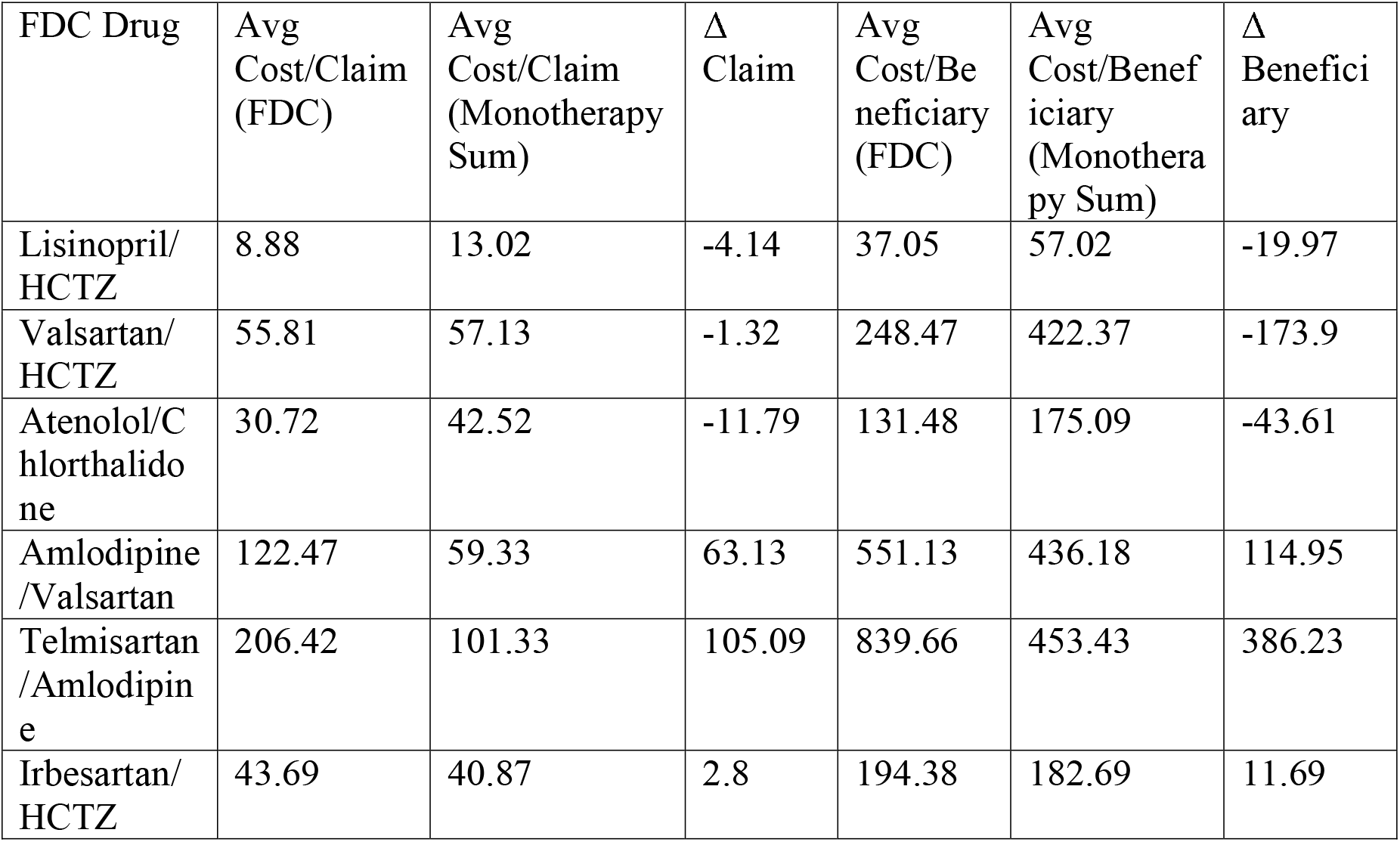
Ten-Year Average Cost Comparison of Fixed-Dose Combination Antihypertensives and Their Monotherapy Components (2013–2022) Average cost per claim and per beneficiary for each fixed-dose combination (FDC) compared to the summed costs of their individual monotherapy components, based on Medicare Part D data. Positive Δ values indicate higher costs for the FDC; negative Δ values indicate FDC cost savings compared to summed monotherapy. Costs are presented in U.S. dollars.

| FDC Drug | Avg Cost/Claim (FDC) | Avg Cost/Claim (Monotherapy Sum) | $\Delta$ Claim | Avg Cost/Beneficiary (FDC) | Avg Cost/Beneficiary (Monotherapy Sum) | $\Delta$ Beneficiary |
| --- | --- | --- | --- | --- | --- | --- |
| Lisinopril/HCTZ | 8.88 | 13.02 | -4.14 | 37.05 | 57.02 | -19.97 |
| Valsartan/HCTZ | 55.81 | 57.13 | -1.32 | 248.47 | 422.37 | -173.9 |
| Atenolol/Chlorthalidone | 30.72 | 42.52 | -11.79 | 131.48 | 175.09 | -43.61 |
| Amlodipine/Valsartan | 122.47 | 59.33 | 63.13 | 551.13 | 436.18 | 114.95 |
| Telmisartan/Amlodipine | 206.42 | 101.33 | 105.09 | 839.66 | 453.43 | 386.23 |
| Irbesartan/HCTZ | 43.69 | 40.87 | 2.8 | 194.38 | 182.69 | 11.69 |

## Discussion

In this national analysis of Medicare Part D prescription data, half of the two-drug fixed-dose combination (FDC) antihypertensive therapies evaluated were less expensive per claim and per beneficiary than prescribing their component drugs separately. These findings indicate that FDC therapies do not uniformly increase costs and that several generic combinations may provide both clinical and economic advantages. Prior clinical trials such as UMPIRE and TRIUMPH [1,2] have shown that FDCs improve adherence and blood pressure control through simplified regimens. Our real-world analysis complements these findings, demonstrating that certain established combinations are cost-efficient in practice and align with the goals of value-based healthcare [5,6,9].

We observed meaningful variation in cost outcomes among the six FDCs analyzed. Combinations containing both an angiotensin II receptor blocker and a calcium-channel blocker, amlodipine/valsartan and telmisartan/amlodipine, had the greatest average cost increases, likely reflecting brand pricing or formulary status [10]. Conversely, valsartan/hydrochlorothiazide demonstrated substantial cost reductions, with more modest but still relevant savings observed for lisinopril/hydrochlorothiazide and atenolol/chlorthalidone. These findings suggest that long-standing generic combinations can offer effective and affordable treatment options that simplify hypertension management. Clinicians should remain mindful of hydrochlorothiazide’s limitations in older adults, particularly its potential for electrolyte imbalance and volume depletion, which may increase fall risk.

From a health-systems standpoint, these results may guide formulary planning and payer policy. Although major hypertension guidelines endorse combination therapy to improve adherence, uptake has remained inconsistent, partly due to cost concerns [3,5,6]. Our analysis suggests that such concerns may be overstated for several generic combinations. Insurers and pharmacy benefit managers could use these findings to support formulary placement or value-based contracting that favors cost-efficient FDCs [8,9]. Broader use of affordable combinations could be particularly valuable for older adults who often face complex regimens and financial barriers [7–9]. Cost trends should be interpreted in the context of clinical effectiveness and adherence. As efforts continue to improve hypertension control and manage healthcare spending, selectively increasing the use of lower-cost, evidence-based FDCs represents a practical and sustainable approach to enhancing the value of cardiovascular care.

## Limitations

This study has several limitations. Medicare Part D data are aggregate and do not include patient-level information such as adherence or dosing. Reported costs reflect total spending and exclude rebates or discounts, so actual payer expenses may differ. The analysis assumed therapeutic equivalence between FDCs and their component drugs, though individual prescribing decisions can differ. Only six two-drug combinations with complete ten-year data were included, and the study focused on descriptive comparisons rather than adjusted statistical modeling. Three-drug combinations, agents with incomplete data, and newer therapies were not analyzed. Some commonly prescribed FDC antihypertensive agents were excluded because their component data were incomplete or inconsistently reported during 2013–2022. Finally, because the dataset reflects Medicare beneficiaries, the findings may not apply directly to younger or privately insured populations with different cost-sharing structures.

## Conclusion

In this national analysis, half of the evaluated fixed-dose combination (FDC) antihypertensive therapies were less costly than prescribing their component drugs separately. These findings challenge the assumption that FDCs inherently carry a financial premium and show that several combinations can provide both adherence advantages and meaningful cost reductions. Valsartan/hydrochlorothiazide and lisinopril/hydrochlorothiazide, in particular, appeared to be cost-efficient and widely used options that warrant consideration in routine practice. Prior clinical trials such as UMPIRE and TRIUMPH have already demonstrated the adherence and blood pressure benefits of FDC therapy. Our analysis complements this evidence by offering real-world economic support for their selective use as part of efforts to enhance value and outcomes in cardiovascular care.

## Data Availability

The study used ONLY openly available human data that were originally located at:
https://data.cms.gov/provider-summary-by-type-of-service/medicare-part-d-prescribers/medicare-part-d-prescribers-by-provider-and-drug/data/2022
Data for earlier years (2013 to 2021) are available through the same CMS portal using the year dropdown menu.

## Data Availability Statement

The Medicare Part D Prescription Drug Event data used in this study are publicly available from the Centers for Medicare & Medicaid Services (CMS) at https://data.cms.gov. All data analyzed were aggregate and de-identified, requiring no institutional review board approval. Specific datasets can be accessed through the “Medicare Part D Drug Spending” section on the CMS website.

## Code Availability

Relevant code used for statistical analysis can be made available upon direct request to the study authors.

## Acknowledgements

The author completed this analysis independently and did not receive external funding or institutional support. Thanks to colleagues who provided helpful feedback on the manuscript during preparation.

## Ethical Approval

This study used publicly available, aggregate Medicare Part D data and did not involve human subjects or identifiable patient information. As such, ethical approval was not required.

